# Genome-wide investigation highlights global and local pleiotropy linking neurodevelopmental disorders to acquired hearing problems

**DOI:** 10.1101/2025.11.03.25339410

**Authors:** Qishu Zhang, Brenda Cabrera-Mendoza, Qianyu Chen, David Davtian, Dan Qiu, Jun He, Renato Polimanti

## Abstract

**Background:** Neurodevelopmental disorders have been associated with hearing problems (HP) later in life, but there is limited information regarding their shared biology.

**Methods:** We leveraged large-scale genome-wide datasets to estimate genetic correlation (global and local), polygenic overlap, and locus-specific pleiotropy among HP, autism spectrum disorder (ASD), attention-deficit/hyperactivity disorder (ADHD), and Tourette syndrome (TS). Then, we investigated shared molecular functions, biological processes, and cellular components, and performed a drug-repurposing analysis to identify compounds that may target the pathogenic processes linking neurodevelopmental disorders to HP.

**Results:** We observed high genetic correlation of HP with ASD (rg=0.72) and TS (rg=0.69). While HP-ADHD genetic correlation was low (rg=0.07), 40% of the causal variants were shared between these conditions, with only 53% of them showing concordant effect directions. We also identified nine chromosomal regions with evidence of ADHD-HP local genetic correlations with pleiotropic effects on other outcomes such as smoking initiation, brain-imaging phenotypes, and bilirubin levels. With respect to HP-ASD, we observed an inverse local genetic correlation within *CD33* chromosomal region. Pleiotropy among HP, ASD, and ADHD was also identified in two variants (rs325485 and rs2207286) included within 95% credible sets related to neuropsychiatric conditions, altered hearing function, and other traits such as risk taking and insomnia. Drug-repurposing analyses identified anisomycin for HP-ASD shared biological mechanisms and five compounds related to HP-ADHD pleiotropy.

**Conclusions:** Our findings provide evidence that the comorbidity between neurodevelopmental disorders and HP is at least partially due to shared pathogenic processes acting through intrinsic and extrinsic factors.

## INTRODUCTION

Hearing problems (HP), including hearing loss, auditory processing deficits, and other related impairments, are among the most common chronic health conditions across lifespan(Alanazi, 2023; Volter *et al*., 2020). Recent studies have highlighted an increased prevalence of acquired HP among individuals affected by neurodevelopmental disorders, such as autism spectrum disorder (ASD), attention-deficit/hyperactivity disorder (ADHD), and Tourette syndrome (TS)(Gunturu *et al*., 2025; Lowick & Mbatha, 2025; McKenna *et al*., 2024). This epidemiological evidence supports possible shared pathogenesis linking neurodevelopmental disorders to altered auditory function later in life. In this context, the loss of *SCN2A* gene in oligodendrocytes has been associated with the disruption of myelin plasticity and neural circuit integrity as well as subsequent alterations in axonal and synaptic function, resulting in auditory processing abnormalities among individuals with ASD(Bae *et al*., 2025).

Additionally, other studies reported that sensory hypersensitivity, including auditory sensitivity, is significantly associated with autistic traits, with attention switching and communication difficulties serving as key predictors(Keshavarz & Esmaeilpour, 2025).

To date, most studies regarding the comorbidity between neurodevelopmental disorders and acquired HP primarily focused on specific biological pathways and molecular targets(McKenna *et al*., 2024; Omidvar *et al*., 2020; Worley *et al*., 2011). Large-scale genome-wide association studies (GWAS) permitted investigators to generate novel insights into the pathogenesis of human traits and diseases(Abdellaoui *et al*., 2023), including neurodevelopmental disorders(Demontis *et al*., 2023; Grove *et al*., 2019; Yu *et al*., 2019) and acquired HP(De Angelis *et al*., 2023). Integrating genome-wide information with multi-modal magnetic resonance imaging data, a brain-wide investigation demonstrated that structural and functional changes in the nervous system can lead to acquired HP in adults(He *et al*., 2024). In this context, a systematic investigation of the pleiotropy linking neurodevelopmental disorders with hearing problems can provide deeper insight into the shared pathogenesis of these conditions and may reveal opportunities for therapeutic interventions that could improve the quality of life of the affected individuals. To our knowledge, although genome-wide data are available for these disorders(De Angelis *et al*., 2023; Demontis *et al*., 2023; Grove *et al*., 2019; Yu *et al*., 2019), no study has fully leveraged them to dissect their genetic overlap. To fill this gap, we applied a multi-layered analytic framework to disentangle HP pleiotropy with ASD, ADHD, and TS. Our analyses investigated both global and local pleiotropy using complementary approaches. Then, we translated our genetic findings into their potential functional implications through gene ontology (GO) and drug-repurposing analyses. Overall, our study demonstrated that the comorbidity between HPs and neurodevelopmental disorders is at least partially due to widespread pleiotropy between these conditions. These findings could be translated into instruments for the early identification of individuals at high comorbidity risk and targets to develop tailored therapeutic strategies.

## MATERIALS AND METHODS

We conducted a genome-wide investigation to assess global and local pleiotropy between HP and multiple neurodevelopmental disorders (ASD, ADHD, and TS). Owing to the use of previously collected, deidentified, and aggregated data, this study did not require institutional review board approval. Ethical approval and written informed consent had been obtained in all original cohorts that enrolled the participants investigated(De Angelis *et al*., 2023; Demontis *et al*., 2023; Grove *et al*., 2019; Yu *et al*., 2019).

### Genome-wide Datasets

We leveraged genome-wide association statistics generated from large-scale studies. Specifically, GWAS of neurodevelopmental disorders were obtained from the Psychiatric Genomics Consortium (available at https://www.med.unc.edu/pgc/download-results/). ASD genome-wide information was derived from a meta-analysis including 18,381 ASD cases and 27,969 controls from clinical cohorts and population-based samples(Grove *et al*., 2019).

ADHD GWAS meta-analysis included 10,608 cases and 225,534 controls from cohorts enrolling participants with different ages of onset(Demontis *et al*., 2023). PGC GWAS of TS was based on 4,819 cases and 9,488 controls(Yu *et al*., 2019). With respect to HP, we leveraged genome-wide association statistics derived from 501,825 adult participants (29% cases) self-reporting information regarding a HP(De Angelis *et al*., 2023) (available at https://zenodo.org/records/7897038). To focus only on acquired HPs, this study excluded cases of congenital HP forms. Due to the limited availability of cohorts representing global populations, the datasets investigated included only individuals of European descent (EUR).

### Global Genetic Correlation and Polygenic Overlap

We applied linkage disequilibrium score regression (LDSC, version 1.0.1; available at https://github.com/bulik/ldsc) to estimate SNP-based heritability and global genetic correlations among HP, ASD, ADHD, and TS. LDSC analysis was conducted using HapMap variants and pre-computed LD scores from 1000 Genomes Project EUR populations. To further investigate HP polygenic overlap with neurodevelopmental disorders, we used the Mixture of eXperts Regression (MiXeR, version 1.2.0).(Frei *et al*., 2019) All analyses were performed using the MiXeR Python wrapper (available at https://github.com/precimed/mixer), with parameter optimization performed via grid search with log-likelihood maximization.

Similarly to LDSC analysis, this analysis was also based on HapMap variants and LD structure of 1000 Genomes Project EUR populations. The bivariate MiXeR analysis was performed to estimate the number of shared causal SNPs and the degree of genetic overlap between traits, beyond what is captured by genetic correlation. Specifically, we estimated the proportion of shared polygenic signal and the concordance ratios indicating the proportion of shared SNPs with concordant effect directions. To define statistically meaningful bivariate MixeR models, we considered only those with Akaike information criterion (AIC) for the best model versus the model with minimal polygenic overlap greater than zero.

### Local Genetic Correlation

We applied Local Analysis of Variant Association (LAVA) approach (version 1.8.0)(Werme *et al*., 2022) to investigate locus-specific pleiotropy between HP and neurodevelopmental disorders. Following LAVA documentation (available at https://github.com/josefin-werme/LAVA), we used 2,495 pre-defined, approximately independent LD blocks from 1000 Genomes Project EUR reference populations. For each trait, we estimate the locus-specific SNP-based heritability. Then, we performed LAVA bivariate analysis to estimate locus-specific genetic correlation between HP and neurodevelopmental disorders. False discovery rate (FDR q<0.05) was applied to define statistically significant local genetic correlations after accounting for the number of loci tested.

### Pleiotropic Loci

To identify individual loci with pleiotropic effects on both HP and neurodevelopmental disorders, we used PolarMorphism approach (available at https://github.com/UMCUGenetics/PolarMorphism).(von Berg *et al*., 2022) This method analyzes genome-wide data to identify genetic variants associated with multiple traits due to horizontal pleiotropy, attenuating the effect of vertical pleiotropy observed in the genetic correlation via a decorrelating transform.(von Berg *et al*., 2022) For each SNP, PolarMorphism computes two parameters: the overall strength of association across traits (r) and the relative contribution of the effect across the traits (theta). FDR was used to define pleiotropic loci after multiple testing correction. PLINK 1.9 (available at https://www.cog-genomics.org/plink/1.9/) was used to identify LD-independent pleiotropic loci, considering a 250-kb window and an LD r²<0.1 based on 1000 Genomes Project Phase 3 EUR reference populations. Functional annotation was performed using Open Target Platform(Buniello *et al*., 2025) (available at https://platform.opentargets.org/) and GTEx v10(GTEx Consortium, 2020) (available at https://www.gtexportal.org/home/).

### Gene Ontology and Drug Repurposing Analyses

To explore the biological processes, molecular functions, and cellular components shared between HP and neurodevelopmental disorders, we applied GSA-MiXeR to estimate gene-set heritability enrichments across these traits.(Frei *et al*., 2024) Specifically, GSA-MiXeR identified GO terms that were significantly enriched among SNPs with shared polygenic signal between two traits. GSA-MiXeR package is available at https://github.com/precimed/gsa-mixer. GO enrichments were assessed using a non-parametric empirical approach, comparing observed enrichment scores against a null distribution generated through randomization.

To define GO terms shared between HP and neurodevelopmental disorders, we applied a Bonferroni multiple testing correction accounting for the number of GOs tested (p<6.31×10^-7^), considering both pair-wise combinations (GO terms that were Bonferroni significant with respect to both HP and one of the neurodevelopmental disorders tested) and a four-way model (GO terms that were Bonferroni significant with respect to HP and all three neurodevelopmental disorders tested).

To reduce redundancy among Bonferroni-significant GO terms and facilitate their interpretation, we applied REVIGO(Supek *et al*., 2011), considering a similarity threshold=0.7 and UniProt as the reference database. Non-redundant Bonferroni filtered GO terms were then used to conduct a drug-repurposing analysis. Specifically, we used the Gene2drug approach(Napolitano *et al*., 2018) to identify molecular compounds associated with transcriptomic profiles overlapping with those of the shared GOs between HP and neurodevelopmental disorders. Transcriptomic reference data were derived from the Connectivity Map drug database (1.5M gene expression profiles from ∼5,000 small-molecule compounds, and ∼3,000 genetic reagents)(Lamb *et al*., 2006).

## RESULTS

### Genetic Correlation and Polygenic Overlap

Considering SNP-based heritability z-scores as a proxy of the statistical power of the GWAS investigated, we observed comparable estimates for HP and ADHD (z=18.1 and 21.6, respectively), which were higher than those observed for TS and ASD (z=8.6 and 8.4, respectively). These differences were mostly explained by the effective sample size of the GWAS investigated (see section Genome-wide Datasets in METHODS). Nevertheless, HP genetic correlations were statistically significant for all neurodevelopmental disorders, although they differed in magnitude (Figure 1, Supplementary Table 1). Specifically, HP showed high genetic correlations with both ASD (rg=0.72±0.03) and TS (rg=0.69±0.03), while the HP-ADHD genetic correlation was statistically significant but relatively low (rg= 0.07±0.01). The extent of HP genetic correlations reflected the pleiotropy among the neurodevelopment disorders investigated (Figure 1), with ASD and TS showing a high genetic correlation (rg=0.63±0.02) and ADHD showing lower estimates with both (ASD-ADHD rg=0.13±0.02; TS-ADHD rg=0.12±0.02).

**Figure 1.**
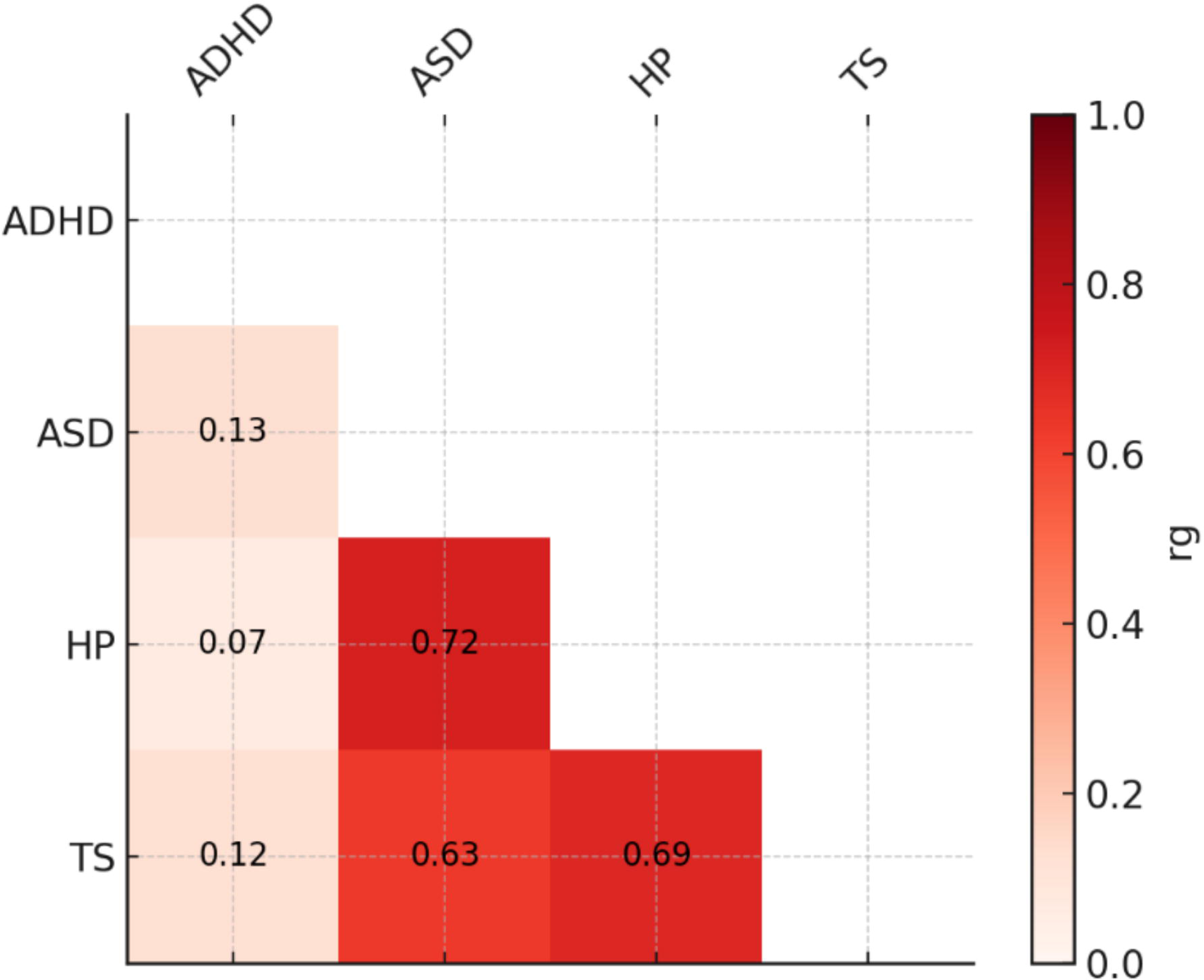
Genetic correlations (rg) among hearing problems (HP), autism spectrum disorder (ASD), attention-deficit/hyperactivity disorder (ADHD), and Tourette syndrome (TS). Color intensity reflects rg magnitude. All rg estimates reported are statistically significant (FDR q<0.05; Supplemental Table 1).

**Figure 2.**
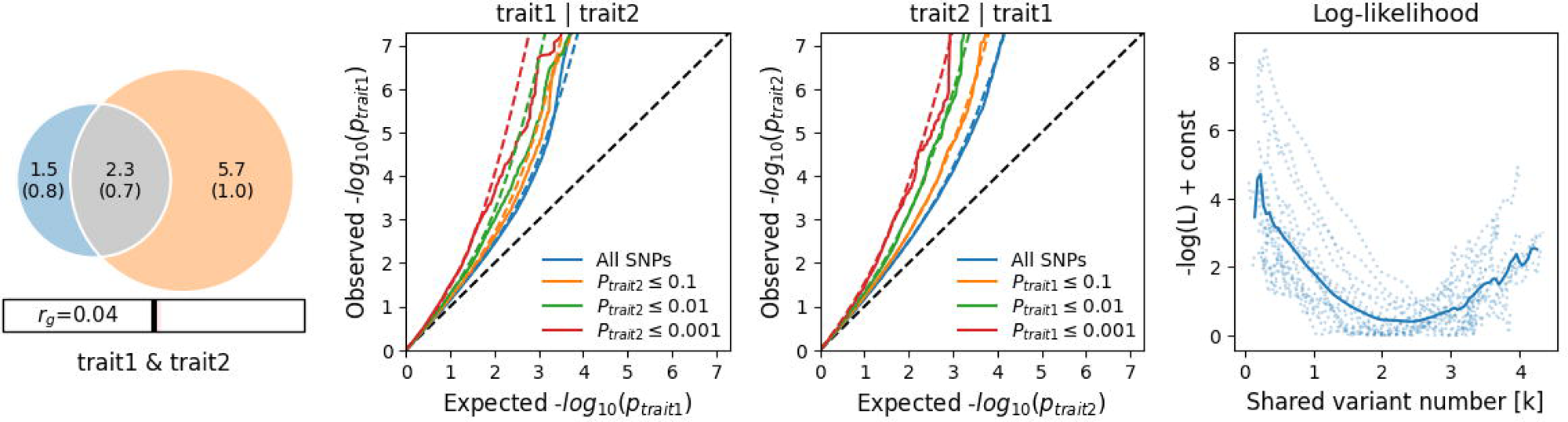
Polygenic overlap between hearing problems (HP, trait1) and attention-deficit/hyperactivity disorder (ADHD, trait2). (a) Venn diagrams of unique and shared polygenic components at the causal level, showing polygenic overlap (gray) between HP (blue) and ADHD (orange). The numbers indicate the estimated quantity of causal variants (in thousands) per component, accounting for 90% of SNP heritability in each phenotype, along with the standard error between parentheses. The size of the circles reflects the degree of polygenicity. (b-c) Conditional Q–Q plots of observed versus expected −log10 p-values in the primary trait as a function of significance of association with a secondary trait at the level of p≤0.1 (orange lines), p≤0.01 (green lines), p≤0.001 (red lines). Blue line indicates all SNPs. Dotted lines in blue, orange, green, and red indicate model predictions for each stratum. The black dotted line represents the expected Q–Q plot under the null hypothesis. (d) Log-likelihood profile for the estimated number of shared causal variants (k). The solid line represents the smoothed likelihood curve, while dotted lines indicate chromosome-wise estimates from the MiXeR fit. The minimum denotes the maximum likelihood estimate of k, supporting the presence of a finite set of shared causal variants.

We observed good fitting statistics only for the HP-ADHD bivariate MiXeR model (AIC_bestVSmininal_=6.13; Figure 1, Supplementary Table 2). As mentioned above, ASD and TS GWAS had limited statistical power due to their smaller effective sample size. While the HP-ADHD genetic correlation was limited (LDSC rg=0.07, MiXeR rg=0.04), 40±11% of the causal variants related to these traits were shared. Within these pleiotropic loci, only 53±2% showed concordant effect directions, and the correlation of effect sizes within HP-ADHD shared polygenic component was 0.11±0.06.

### Local Genetic Correlation

We identified ten FDR-significant locus-specific genetic correlations, with nine of them related to HP-ADHD and one to HP-ASD (Table 1). There was no overlap between the FDR-significant local genetic correlations related to HP-ADHD and HP-ASD. Only the HP-ADHD local genetic correlation on chromosome 12 (hg19 position: 116,197,676-117,091,843; rho=0.76, p=1.16X10^-5^, FDR q=0.008) showed a nominal HP-ASD replication (p=0.017). Within this chromosomal region, a total of 127 genome-wide associations were reported in the GWAS Catalog(Sollis *et al*., 2023) (Supplemental Table 3), including variants associated with skeletal phenotypes (e.g., bone mineral density), immune-related traits (e.g., acute myeloid leukemia and monocyte count), and cardiovascular outcomes (e.g., blood pressure and atrial fibrillation). With respect to HP, only genome-wide significant gene-gene interaction was reported (rs4301303-rs10444487 p=7X10^-9^). While no association was reported with respect to ADHD, ASD, or other psychiatric disorders, there was evidence related to behavioral traits (e.g., educational attainment, rs76130193 p=8X10^-18^; smoking initiation, rs71469748 p=4X10^-11^) and brain-imaging phenotypes (e., vertex-wise sulcal depth rs74550975 p=9X10^-10^; cortical surface area rs74550975 p=4X10^-11^). Interactive effects between smoking status and diastolic pressure were also identified within this chromosomal region (rs11067762 p=5X10^-18^).

**Table 1.** Local genetic correlations between HP and Neurodevelopmental Disorders (autism spectrum disorder, ASD; attention-deficit hyperactivity disorder) surviving false discovery rate multiple testing correction (FDR q < 0.05). NS: not significant; BP: base pairs.

| Chr | Start (BP) | Stop (BP) | Phenotype | rho | p | FDR | GWAS Catalog associations, N |
| --- | --- | --- | --- | --- | --- | --- | --- |
| 2 | 9,960,671 | 10,611,654 | ADHD | 0.911 | 2.39E-04 | 0.047 | 123 |
| 2 | 234,115,093 | 234,945,577 | ADHD | 0.976 | 2.01E-04 | 0.044 | 1,141 |
| 4 | 79,880,102 | 81,206,182 | ADHD | 1.000 | 1.86E-05 | 0.009 | 565 |
| 4 | 102,544,804 | 104,384,534 | ADHD | 1.000 | 5.39E-05 | 0.018 | 1,935 |
|  |  |  | ASD | 0.102 | 0.773 | NS |  |
| 6 | 149,278,799 | 150,635,315 | ADHD | 1.000 | 1.15E-06 | 0.002 | 243 |
|  |  |  | ASD | 0.141 | 0.580 | NS |  |
| 9 | 37,687,113 | 38,822,977 | ADHD | 0.838 | 4.08E-06 | 0.004 | 129 |
| 12 | 116,197,676 | 117,091,843 | ADHD | 0.759 | 1.16E-05 | 0.008 | 127 |
|  |  |  | ASD | 0.481 | 0.017 | NS |  |
| 17 | 27,344,402 | 29,783,141 | ADHD | 1.000 | 1.13E-04 | 0.028 | 1,018 |
| 19 | 51,259,179 | 51,903,804 | ASD | -0.712 | 9.81E-05 | 0.028 | 786 |
|  |  |  | ADHD | -0.302 | 0.209 | NS |  |
| 20 | 58,780,895 | 59,560,457 | ADHD | 0.869 | 5.12E-05 | 0.018 | 27 |
|  |  |  | ASD | -0.366 | 0.247 | NS |  |

Among the other loci specific to HP-ADHD pleiotropy, the most significant result was on chromosome 6 (hg19 position: 149,278,799-150,635,315; rho=1, p=1.15X10^-6^, FDR q=0.002). More than 240 genome-wide significant associations were reported within this chromosomal region in the GWAS catalog (Supplemental Table 4). The most significant included retinal thickness (rs57215159 p=5×10^-72^), atrial fibrillation (rs117984853 p=4×10^-55^), and several brain-imaging phenotypes (e.g., brain morphology rs7752089 p=2×10^-31^; subcortical volume rs12523793 p=2×10^-24^; vertex-wise sulcal depth rs14314 p=7×10^-17^).

Some HP-ADHD pleiotropic regions (i.e., hg19 chr2:9,960,671-10,611,654; chr4:79,880,102-81,206,182; chr9:37,687,113-38,822,977; chr17:27,344,402-29,783,141; chr20:58,780,895-59,560,457) showed genome-wide significant associations that converged on phenotypic traits, such as bone mineral density, blood pressure, atrial fibrillation, hematologic biomarkers, smoking initiation, and educational attainment (Supplemental Tables 5-9). Conversely, other HP-ADHD pleiotropic regions showed different association patterns. For example, within hg19 chr2:234,115,093-234,945,577 region (Supplemental Table 10), we identified the locus most strongly associated with bilirubin levels (e.g., rs6704644 p=1×10^-1043^). Within hg19 chr4:102,544,804-104,384,534 region, multiple genome-wide significant associations were reported for brain iron levels (e.g., pallidum iron levels rs13107325 p=2×10^-231^; substantia nigra iron levels rs13107325 p=2×10^-197^; caudate iron levels rs13107325 p=2×10^-167^) along with several brain-imaging phenotypes (Supplemental Table 11). The only HP-ASD pleiotropic locus identified (hg19 chr19:51,259,179-51,903,804) showed an inverse local genetic correlation (rho=-0.71, p=9.81×10^-5^, FDR q=0.028) and included many genome-wide significant associations with immune-related phenotypes (Supplemental Table 11), with those related to CD33 levels among the strongest ones (e.g., myeloid cell surface antigen CD33 levels rs33978622 p=1×10^-223^).

### Pleiotropy analysis

Our genome-wide pleiotropy analysis using PolarMorphism identified specific variants showing horizontal pleiotropy (shared pathogenic effects) between HP and neurodevelopmental disorders. Because of the limited GWAS sample size for single-variant discovery, TS was excluded from this analysis. Modeling pleiotropic effects among HP, ASD, and ADHD, we identified two LD-independent variants with FDR-significant r and theta statistics, which correspond to the overall strength of association across traits and the relative contribution of the effect across the traits, respectively (Supplemental Table 13). On chromosome 5, rs325485*A allele showed genome-wide significant association with both HP (beta=0.026, p=2.57×10^-8^) and ADHD (beta=0.067, p=2.74×10^-12^) and consistent effect direction on ASD (beta=0.081, p=0.005). According to Open Target Platform(Buniello *et al*., 2025), this variant is included within 57 95%-credible sets related to a range of complex traits (Supplemental Table 14). The top associations included hand grip strength, ASD, depression medications, risk-taking tendency, and asthma-depression comorbidity. Additionally, rs325485 is both an expression quantitative trait locus (QTL) and a splicing QTL for *ENSG00000251574* in testis (Supplemental Table 15). The other pleiotropic locus was rs2207286 on chromosome 1, which showed concordant effects across HP (beta=0.019, p=1.7×10^-4^), ASD (beta=0.096, p=0.001), and ADHD (beta=0.036, p=2.86×10^-4^). This variant is included within 11 95%-credible sets available from the Open Target Platform(Buniello *et al*., 2025), with the most significant ones related to insomnia, middle ear disease, acquired hearing loss, general risk tolerance, and non-suppurative otitis media (Supplemental Table 16).

### Gene Enrichment and Drug-Repurposing Analyses

Through GSA-MiXeR approach (Frei *et al*., 2024), we identified 56 GO terms that reached Bonferroni significance with respect to both HP and all neurodevelopmental disorders investigated (p<6.31×10^-7^; Figure 3, Supplemental Table 17). Among these, “*plasma membrane region*” (GO:0098590) was the most significant GO term for both ASD (p=9.87×10^-52^) and TS (p=3.94×10^-47^), while “*supramolecular complex*” (GO: GO:0099080) was the most significant for both HP (p=1.51×10^-64^) and ADHD (p=3.25×10^-37^). For the subsequent drug-repurposing analysis, we considered both Bonferroni-significant GO terms shared across all four traits investigated and those shared between HP and each neurodevelopmental disorder tested. With respect to GO terms shared between HP and ASD, we identified a significant enrichment for the transcriptomic profile related to anisomycin (ES=−0.54, p=1.93×10□□). Conversely, HP-ADHD shared GO terms were related to five molecular compounds: perphenazine (ES=0.69, p = 3.11×10□□), bufexamac (ES=0.74, p=5.41×10□□), S-propranolol (ES=0.67, p=6.87×10□□), trichostatin A (ES=0.73, p=6.98×10□□), and propylthiouracil (ES=0.73, p=8.24×10□□).

**Figure 3.**
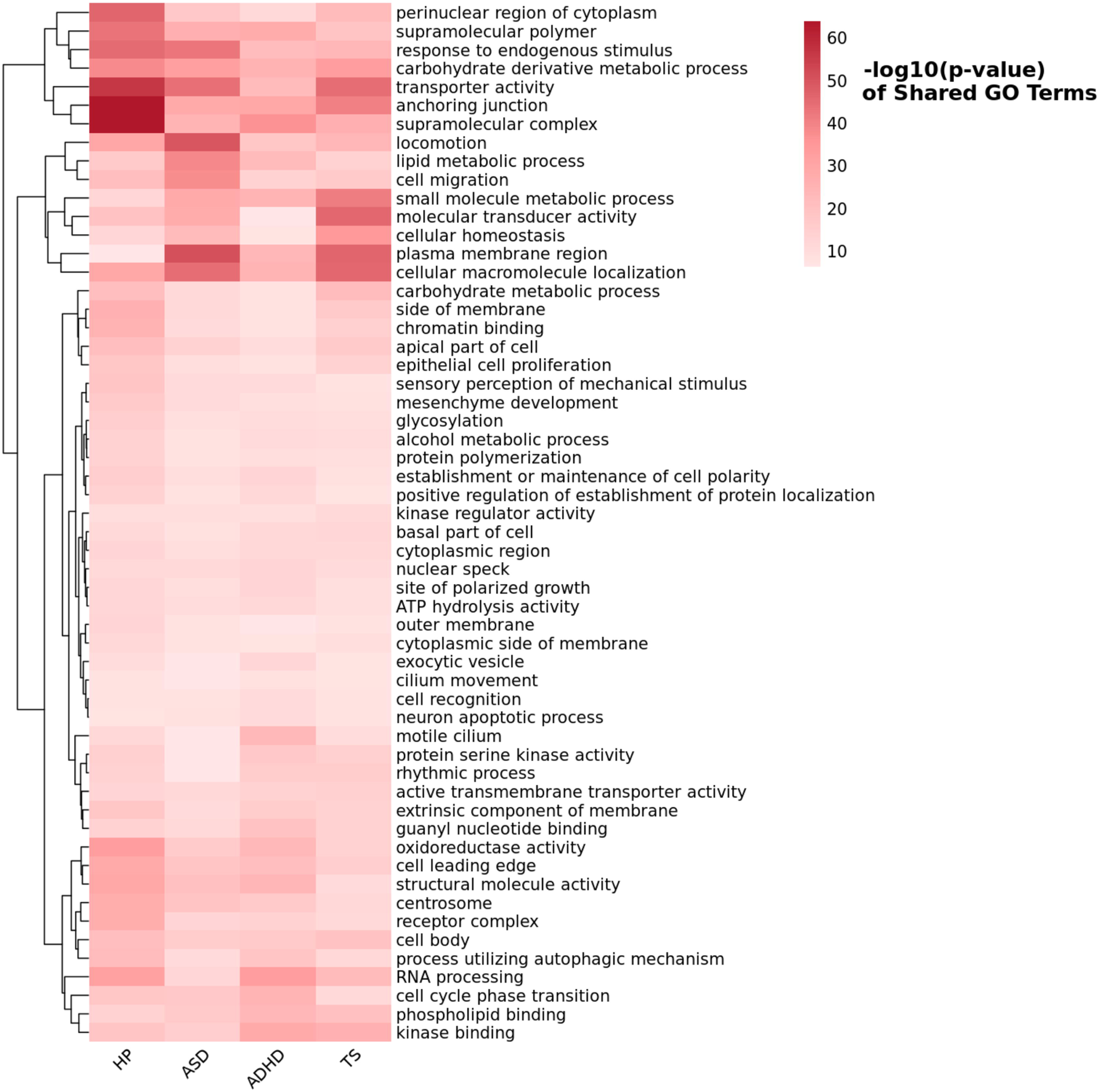
Heatmap of GO Terms reaching Bonferroni significance (p<6.31×10^-7^) shared among hearing problems (HP), autism spectrum disorder (ASD), attention deficit/hyperactivity disorder (ADHD), and Tourette syndrome (TS). The color scale represents the −log (p-value) of each GO term, with deeper red indicating stronger statistical significance.

## DISCUSSION

While extensive evidence exists regarding the comorbidity and the underlying pathogenic processes linking neurodevelopmental disorders and congenital hearing impairments (Akrich *et al*., 2023), little is known regarding the dynamics contributing to the relationship observed between neurodevelopmental disorders and acquired HP(Danyluk & Jacob, 2023).

Leveraging large-scale genome-wide datasets, the present study uncovered global and local pleiotropy between three neurodevelopmental disorders (ASD, ADHD, and TS) and HP, generating new insights into the underlying pathogenetic processes and potential molecular targets involved in this comorbidity. Our global genetic correlation analysis highlighted different degrees of pleiotropy between HP and the neurodevelopmental disorders investigated. Specifically, ASD and TS showed much larger genetic correlations with HP (rg>0.65) than ADHD (rg<0.1). Clinical and epidemiological studies have reported that individuals affected by ASD are more likely to be affected by hearing impairment over their lifetime than the general population and age-matched population groups(Demopoulos & Lewine, 2016; Nian *et al*., 2024). Conversely, no information is available regarding the relationship between TS and HP.

Although HP-ADHD genetic correlation was much lower than that observed for ASD and TS, a nationwide, population-based, cross-sectional study of adolescents reported a strong association between hearing impairment and severe ADHD with consistent relationships across hearing impairment type (sensorineural vs. conductive) and severity(Tsur *et al*., 2024).

These previous findings are in line with HP-ADHD polygenic overlap identified by our MiXer analysis. Indeed, while the HP-ADHD genetic correlation was limited (rg<0.1), 40% of causal variants were shared between these conditions, with >50% of them showing concordant effect directions. Although HP and ADHD showed a limited global genetic correlation, our findings suggest that individual loci shared between these conditions may contribute to their comorbidity. Indeed, we identified 10 chromosomal regions with strong evidence of local genetic correlation. Several of these have been previously associated with traits that may play a role in HP-ADHD co-occurrence. For instance, some of the loci identified were linked to brain structural changes, which may connect previous findings on neuroimaging differences in ADHD cases(Lohani & Rana, 2023; Pereira-Sanchez & Castellanos, 2021) and recent findings connecting central nervous system alterations to HP(He *et al*., 2024). Additionally, one of the HP-ADHD loci was previously associated with iron levels in multiple brain regions. A systematic review of neuroimaging studies reported significantly reduced brain iron content in medication-naïve children with ADHD(Morandini *et al*., 2024). Similarly, iron deficiency anemia was associated with both sensorineural and combined hearing loss in adults(Schieffer *et al*., 2017). These findings suggest that altered iron regulation may play a role in ADHD-HP shared pathogenesis.

Two other HP-ADHD loci supported additional pathogenic mechanisms. On chr 2 (hg19 position: 234,115,093-234,945,577), HP-ADHD local genetic correlation overlapped with the most significant locus associated with bilirubin levels(Sinnott-Armstrong *et al*., 2021).

Neonatal jaundice (i.e., elevated blood level of bilirubin in newborns) has been associated with ADHD and other neurodevelopmental disorders, although the pathogenic mechanisms remain unclear(Wei *et al*., 2015). In the context of HP, bilirubin-induced neurologic damage has been linked to auditory neuropathy spectrum disorder(Olds & Oghalai, 2015). On chromosome 6 (hg19 position: 149,278,799-150,635,315), HP-ADHD local genetic correlation overlapped with a genome-wide significant association with retinal thickness. A recent study reported that ADHD patients have a significant reduction in total and outer retinal thickness across several macular sectors in both eyes, highlighting retinal thickness as a potential biomarker for this condition(Miquel-Lopez *et al*., 2025). With respect to HP, retinal thickness and related measurements have been investigated as predictors of aging-related functional decline(Lee *et al*., 2025).

Multiple ADHD-HP loci were related to smoking initiation. Extensive literature has reported both earlier smoking initiation among ADHD cases(van Amsterdam *et al*., 2018) and the impact of tobacco smoking on HP later in life(Garcia Morales *et al*., 2022). This could point out smoking initiation as a moderator linking ADHD to HP. Similarly, other phenotypes associated across HP-ADHD loci (i.e., educational attainment, cardiovascular conditions, and immune alterations) that could connect ADHD to HP(Baiduc *et al*., 2023; Stonkute & Vierboom, 2025; Tsur *et al*., 2024; Zhang *et al*., 2024).

Beyond HP-ADHD pleiotropic loci, we also identified a genomic region on chromosome 19 (hg19: 51,259,179-51,903,804), showing an inverse local genetic correlation between HP and ASD. Specifically, while the direction of global and local pleiotropy between HP and ADHD was concordant (positive, i.e. increased ADHD genetic risk was correlated with increased HP genetic risk), we observed a discordant effect direction between HP-ASD global genetic correlation (positive) and HP-ASD local genetic correlation on chromosome 19 (inverse, i.e. increased ASD genetic risk was correlated with reduced HP genetic risk). Within the genomic region on chromosome 19, there are several variants associated with immune-related traits, mainly related to *CD33* gene (Supplemental Table 11), which encodes a transmembrane protein receptor expressed on myeloid cells. Genetically regulated CD33 levels in myeloid cells have been associated with increased odds of ADHD(Jue *et al*., 2024). No information is currently available regarding whether CD33 may be implicated in HP pathogenesis. However, there is an extensive literature regarding CD33 role in Alzheimer’s disease and the different impact of its isoforms on neurodegeneration(Eskandari-Sedighi *et al*., 2023). We hypothesize that the inverse local genetic correlation between HP and ASD observed on chromosome 19 may be related to regulatory mechanisms that differentially influence these conditions.

Supporting this, many CD33 expression QTLs have been identified by GTEx v10(GTEx Consortium, 2020) (available at https://www.gtexportal.org/home/). While most of them showed cross-tissue effects, there were also CD33 expression QTLs in only a single tissue (Supplemental Table 18). Caudate nucleus showed the highest number of tissue-specific CD33 expression QTLs (Supplemental Table 18). Interestingly, this brain region has been linked to both ASD and HP, with increased caudate volume being associated with restricted and repetitive behaviors in ASD cases(Qiu *et al*., 2016) and with sensory gating function of caudate nucleus linked to altered hearing perception(Wang *et al*., 2021).

While our local genetic correlation analysis did not identify genomic regions linking HP to both ADHD and ASD, we identified two single variants with shared genetic effects between HP and these neurodevelopmental disorders. According to Open Target Platform(Buniello *et al*., 2025), these loci are within 95% credible sets for multiple complex traits. With respect to rs325485 on chromosome 5, top associations were related to several mental health phenotypes, including ASD, depression medications, and risk-taking tendency, which support pleiotropy with neurodevelopmental disorders. This variant was also related to asthma-depression comorbidity (Supplemental Table 14), highlighting its potential implication in comorbidities between mental and physical health. In particular, its relationship with asthma may have a direct effect on HP risk, as asthma is associated with an increased risk of sensorineural hearing loss(Choi *et al*., 2022). With respect to rs2207286 on chromosome 1, the top 95% credible sets were related to several HL-related phenotypes in addition to general risk tolerance (a trait well-known to be associated with ADHD(Pollak *et al*., 2023)) and insomnia. The latter is commonly observed in neurodevelopmental disorders(Shelton & Malow, 2021) and HP(Clarke *et al*., 2019), suggesting its potential role in contributing to the pleiotropy observed.

Through the GO analysis, we found a significant enrichment for multiple biological processes, molecular functions, and cellular components in the genetic heritability shared across HP and the three neurodevelopmental disorders investigated. These were mostly related to gene-sets related to broad domains, suggesting that the pathogenesis shared between these conditions converges on mechanisms acting across different biological systems. Nevertheless, when considering gene-sets shared between HP and each of the neurodevelopmental disorders tested, we identified specific molecular compounds showing transcriptomic signatures overlapping with the GOs identified. Specifically, HP-ASD drug-repurposing analysis pointed to anisomycin, an antibiotic that inhibits bacterial protein and DNA synthesis (DrugBank Accession Number: DB07374). This appears to converge with the identification of HP-ASD local genetic correlation observed in *CD33* locus, which plays an important role in innate immunity(Malpass, 2013). With respect to HP and ASD, anisomycin has been associated with impaired social recognition when administered in certain brain regions(Pena *et al*., 2014), and its chronic exposure also appears to induce hair cell death(Yuan *et al*., 2021). ADHD-HP drug-repurposing analysis identified five molecular compounds: perphenazine, trichostatin, propylthiouracil, propranolol, and bufexamac. Perphenazine, a first-generation antipsychotic medication that acts by blocking dopamine receptors in the brain (DrugBank Accession Number DB00850). This drug is also indicated for the treatment of Meniere’s disease, an inner ear disorder characterized by symptoms such as vertigo, tinnitus, and hearing loss(Gupta & Jamwal, 2022). Trichostatin is an inhibitor of histone deacetylase (HDAC) enzymes (DrugBank Accession Number DB04297), which has been reported to reduce hearing loss(Nam *et al*., 2025) and maladaptation-induced emotional abnormality in animal models(Kimijima *et al*., 2022). Recently, the transcriptomic profile of HDAC inhibitors has been linked to the comorbidity among psychiatric disorders, spine degenerative disease, and chronic pain(Qiu *et al*., 2025). Propylthiouracil is an antithyroid medication (DrugBank Accession Number: DB00550) that has been associated with rare cases of sudden hearing loss(Tanabe *et al*., 2021) and is being used to induce hyperactive behaviors related to perinatal hypothyroidism in mice(Umezu *et al*., 2019). Propranolol is a beta-blocker medication used to treat cardiovascular and anxiety-related conditions (DrugBank Accession Number: DB00571). There was some earlier evidence regarding the possible benefits of propanol to treat ADHD symptoms(Mattes, 1986). Bufexamac is a non-steroidal anti-inflammatory drug used to treat inflammatory skin conditions (DrugBank Accession Number: DB13346), but there is no evidence linking this drug to HP or ADHD. These molecular compounds highlight the presence of druggable pathways shared between HP and neurodevelopmental disorders, supporting the potential of identifying therapeutic interventions to reduce their comorbidity.

While we believe our results open new directions in neuropsychiatric research, we also acknowledge two important limitations. First, our analysis was limited to EUR GWAS due to the limited availability of large-scale datasets for other population groups. Because of population differences in both neurodevelopmental disorders and HP(Gallin *et al*., 2025; He *et al*., 2025; Shalaby *et al*., 2024), it will be important to assess the generalizability of our findings across human groups when novel datasets are available. Second, because of different GWAS statistical power, we could not investigate the three neurodevelopmental disorders with the same analytic depth, which led to an imbalance regarding the characterization of HP pleiotropy across conditions tested.

In conclusion, our study demonstrated that the pleiotropy between HP and neurodevelopmental disorders at least partially contributes to the comorbidity observed between these conditions. In particular, our findings highlighted that the relationship between neurodevelopmental disorders and altered hearing function can lead to acquired HP later in life through both intrinsic and extrinsic pathogenic processes.

## DATA AVAILABILITY

The data generated by the present study are included in the manuscript and its supplemental material. Genome-wide association statistics from the Psychiatric Genomics Consortium can be accessed at https://pgc.unc.edu/for-researchers/download-results/. HP GWAS data can be accessed at https://zenodo.org/records/7897038.

## CODE AVAILABILITY

The analyses presented in the current study were conducted using publicly available code. LDSC, https://github.com/bulik/ldsc; MiXeR, https://github.com/precimed/mixer; LAVA, https://github.com/josefin-werme/LAVA; PolarMorphism, https://github.com/UMCUGenetics/PolarMorphism; PLINK, https://www.cog-genomics.org/plink/1.9/; GSA-MiXeR, https://github.com/precimed/gsa-mixer.

## Data Availability

The data generated by the present study are included in the manuscript and its supplemental material.

## ACKNOWLEDGEMENTS

The authors thank the Psychiatric Genomics Consortium for making their data available.

## CONTRIBUTORS

Conceptualization: QZ, RP; Methodology: QZ, BCM, QC, JH, RP; Investigation: QZ, BCM, QC, DD, DQ, JH, RP; Visualization: QZ; Funding acquisition: RP; Supervision: RP; Writing – original draft: QZ, RP; Writing – review & editing: QZ, BCM, QC, DD, DQ, JH, RP.

